# A New Mathematical Approach for the Estimation of epidemic Model Parameters with Demonstration on COVID-19 Pandemic in Libya

**DOI:** 10.1101/2020.07.19.20157115

**Authors:** Mohamed Elmehdi Saleh, Zeinab Elmehdi Saleh

## Abstract

**Background:** The SEIR model or a variation of it is commonly used to study epidemic spread and make predictions on how it evolves. It is used to guide officials in their response to an epidemic. This research demonstrates an effective and simple approach that estimates the parameters of any variations of the SEIR model. This new technique will be demonstrated on the spread of COVID-19 in Libya.

**Methods:** A five compartmental epidemic model is used to model the COVID-19 pandemic in Libya. Two sets of data are needed to evaluate the model parameters, the cumulative number of symptomatic cases and the total number of active cases. This data along with the assumption that the cumulative number of symptomatic cases grows exponentially, to determine most of the model parameters.

**Results:** Libya’s epidemic start-date was estimated as *t*_*o*_ = −18 · 5 days, corresponding to May 5th. We mathematically demonstrated that the number of active cases follows two competing exponential distributions: a positive exponential function, measuring how many new cases are added, and a negative exponential function, measuring how many cases recovered. From this distribution we showed that the average recovery time is 48 days, and the incubation period is 15.2 days. Finally, the productive number was estimated as R_0_ = 7·6.

**Conclusions:** With only the cumulative number of cases and the total number of active cases of COVID19, several important SEIR model parameters can be measured effectively. This approach can be applied for any infectious disease epidemic anywhere in the world.

## INTRODUCTION

Compartmental models such as SEIR is one of the most commonly used model to study epidemic spread ^1–7^ and make predictions on how an epidemic will evolve in the future ^1,4,5,8–11^. The compartmental models lead to a system of nonlinear ordinary differential equations with some parameters that define how the epidemic evolves and how an infected individual moves from one state of the disease to another. The epidemiologist’s goal is to solve for these parameters which is difficult task considering the nature and nonlinearity of the differential equations. Consequently, many researchers rely on empirical values or assume a value for some of the parameters ^1,2,6,7^. These assumptions usually lead to discrepancies in the results or in the prediction of how an epidemic grows ^12^. Recognizing this, we devised a new approach and developed a technique never before used, in order to estimate most of the parameters from the model itself without any assumption of any parameter except that the number of cumulative cases is assumed to follow an exponential growth which is expected in any epidemic. This new approach is general and can be applied on any compartmental model, but for the sake of clarity we demonstrated it here on the spread of COVID-19 in Libya.

In Libya, the development of COVID-19 cases differs than most countries where the epidemic progression can be divided into three stages. As shown in Figure 1. Stage I: The first known case was discovered on March/24/2020, and since then there was a steady increase in the number of cases till April/20/2020 where the number of cases reached 60. Stage II: from April/21/2020 till May/23/2020 the number of total cases was relatively constant throughout this period and there was a slight uptick in the number of cases at the end of this stage where the number of total cases reached 75. Stage III: Since May/24, the number of cases started to exponentially grow and rise rapidly, which alarmed the medical community in Libya.

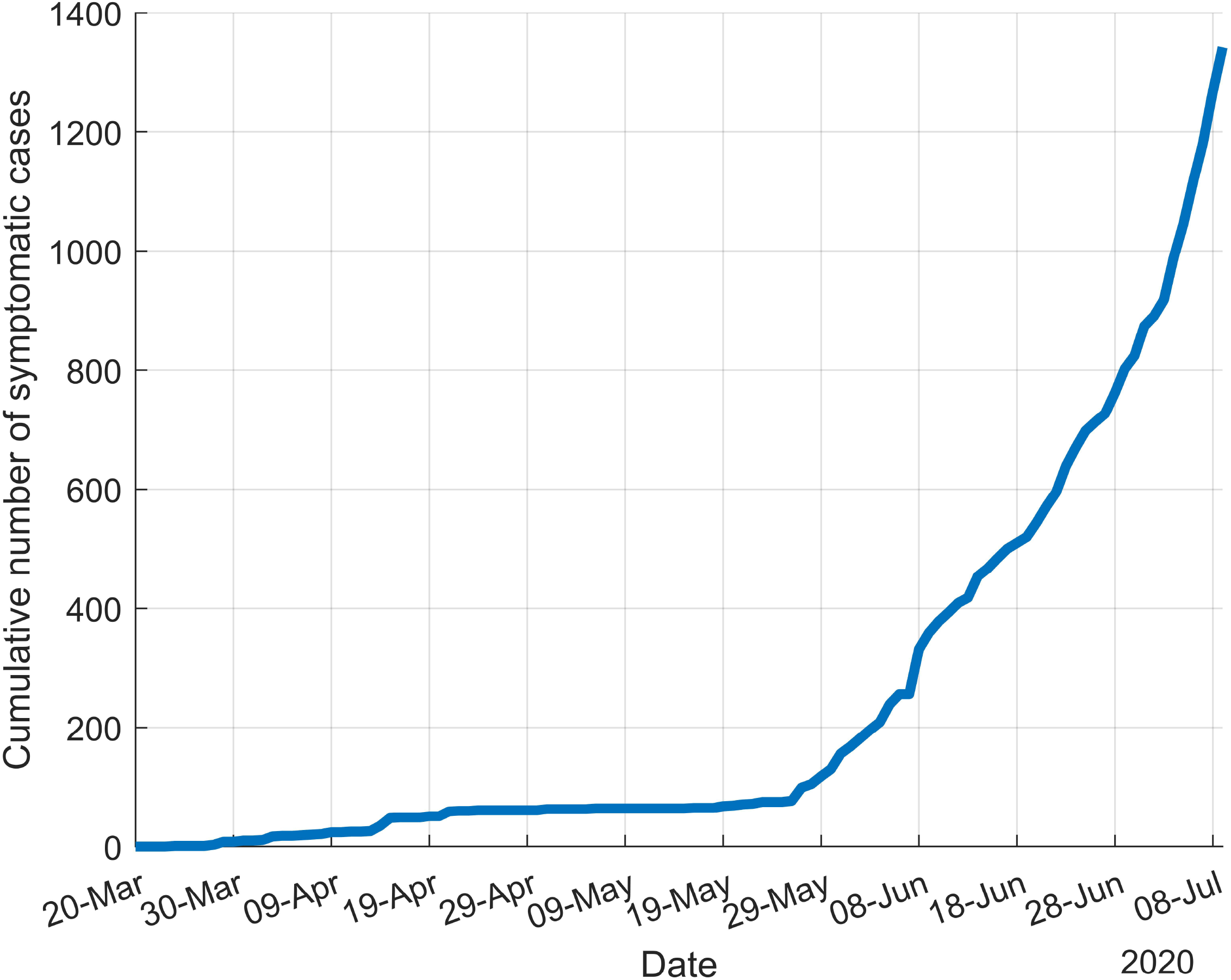

A compartmental model is used to model the epidemic in Libya between the period of May 24^th^ – June 4^th^. Our analysis focused on this period because this is the time when the epidemic started and when it showed an exponential growth pattern. One distinguishing aspect of Covid-19 is that it can be transmitted from infected individuals to healthy ones during the incubation period (pre-symptomatic infection), and it can also be transmitted from an infected individuals who doesn’t show any symptoms (or have mild symptoms) after the incubation period (Asymptomatic infection) ^23,24^. These aspects were included in our model.

## METHODS

The epidemic model as shown in Figure 2 is divided into five compartments and every individual in the population is assumed to be in one of these compartments. Susceptible, S(t): people in this state are healthy but at risk of contracting the virus at any time. Pre-symptomatic infectious, I(t): people who are at the early stages of contracting the virus (Incubation period) and did not shows symptoms yet but can transmit the virus to others. Symptomatic infectious (Active cases), A(t): those who show the sign of virus and are being hospitalized or quarantined therefore, they cannot transmit the virus. Unreported Asymptomatic infectious (Unknown cases) U(t): those individuals who do not show any sign of sickness and do not know that they have the virus, this group can easily transmit the virus to others. Removed state, R(t): those individuals who die or recover from the virus. *C*(*t*)is not an epidemiological state, rather it’s a way to keep track of the cumulative number of active cases from the moment they become symptomatic infectious.

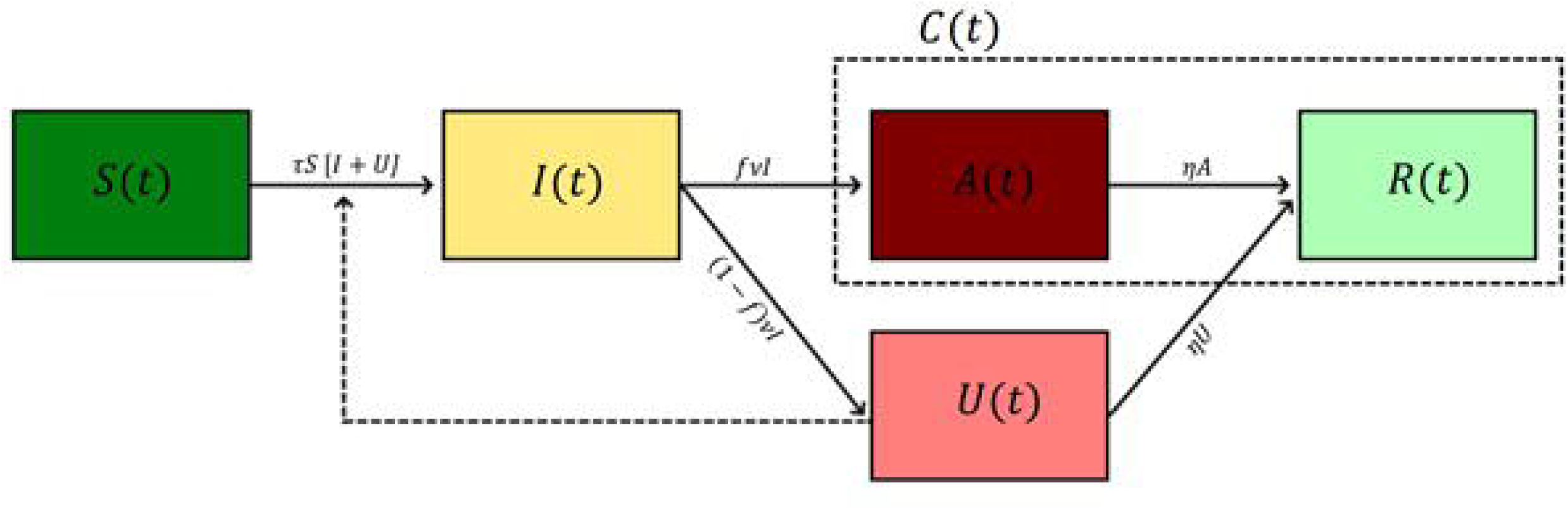

There are two states where the disease can be transmitted from a sick person to a healthy one (Pre-symptomatic infection I(t) and Unreported asymptomatic infectious U(t)) Therefore, a susceptible person in state, S(t) enters the pre-symptomatic infectious class, I(t), at a rate *τ* [*I*(*t*) + *U*(*t*)] where *τ* is the transmission rate. After an incubation period of 1/*υ* a percentage *f* of the pre-Symptomatic individuals develop symptoms or are discovered by the health system, thus, enters the Symptomatic active state A(t) and either enter hospital or quarantine, *f* has a value between 0 and 1. The rest of the pre-symptomatic individuals (1 − *f*) do not show signs of sickness and move to the Asymptomatic unknown state U(t). After a recovery period, both Symptomatic and Asymptomatic individuals move to the recovered state either by death or recovery at the rate *η*. Where 1/*η* is the recovery period.

Also, we will assume the first individual got infected at time *t*_*o*_, so *t*_*o*_ represent the starting time of the epidemic. This person will go through an incubation period 1/*ν*_*o*_ to show any symptoms, therefore he will show symptoms at time *t*_1_. This distinction between the starting time for the epidemic and the starting time when the first active case showed symptoms is really important in solving our model as we will see later. The important parameters are listed in Table-A.1 in the supporting document

The above process can be modeled by the following system of ordinary differential equations.

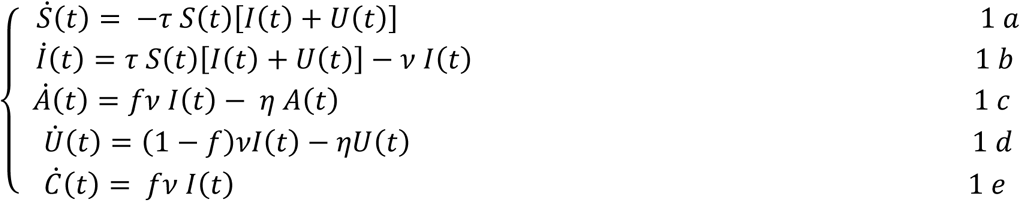

Where the dot indicates derivative with respect to time i.e. 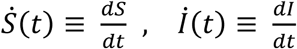‥. etc.

This is a system of nonlinear ordinary differential equations where the exact solution is difficult to find, however, we will show that equation (1c) and (1d) can be reduced to linear ordinary differential equations of the first order which can easily be solved if the number of cumulative cases follows known distribution such as exponential distribution.

## RESULTS

To estimate the model parameters, two sets of reported data are used which are the cumulative number of symptomatic cases and the active number of cases. This data is shown in Table-A.2 in the supporting documents. The process includes fitting the cumulative number of symptomatic cases to exponential function, and then it will be shown that mathematically, the number of active cases has to fit two exponential functions.

### Cumulative number of cases and the starting time of epidemic

Usually, the cumulative number of infected individuals in any epidemic grows exponentially, therefore, it’s customary to assume the cumulative number of cases follows an exponential function and the constants of the function are evaluated from the data. Thus, we will assume the cumulative number of cases follows the form

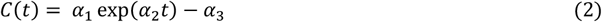

Where *α*_*1*_, *α*_2_ and *α*_*3*_ are constants that needs to be evaluated from the data. For this exponential equation, a least square regression with numerical integration is used to evaluate the constants. The steps for the regression are detailed in Appendix B, while the results are shown in Figure 3. The results for the constants *α*_*1*_, *α*_2_ and *α*_*3*_ are 76.34, 0.0923 and 13.8 respectively. Also, the starting time of the epidemic *t*_*o*_ can be estimated by setting *C*(*t*_*o*_) = 0 which yields

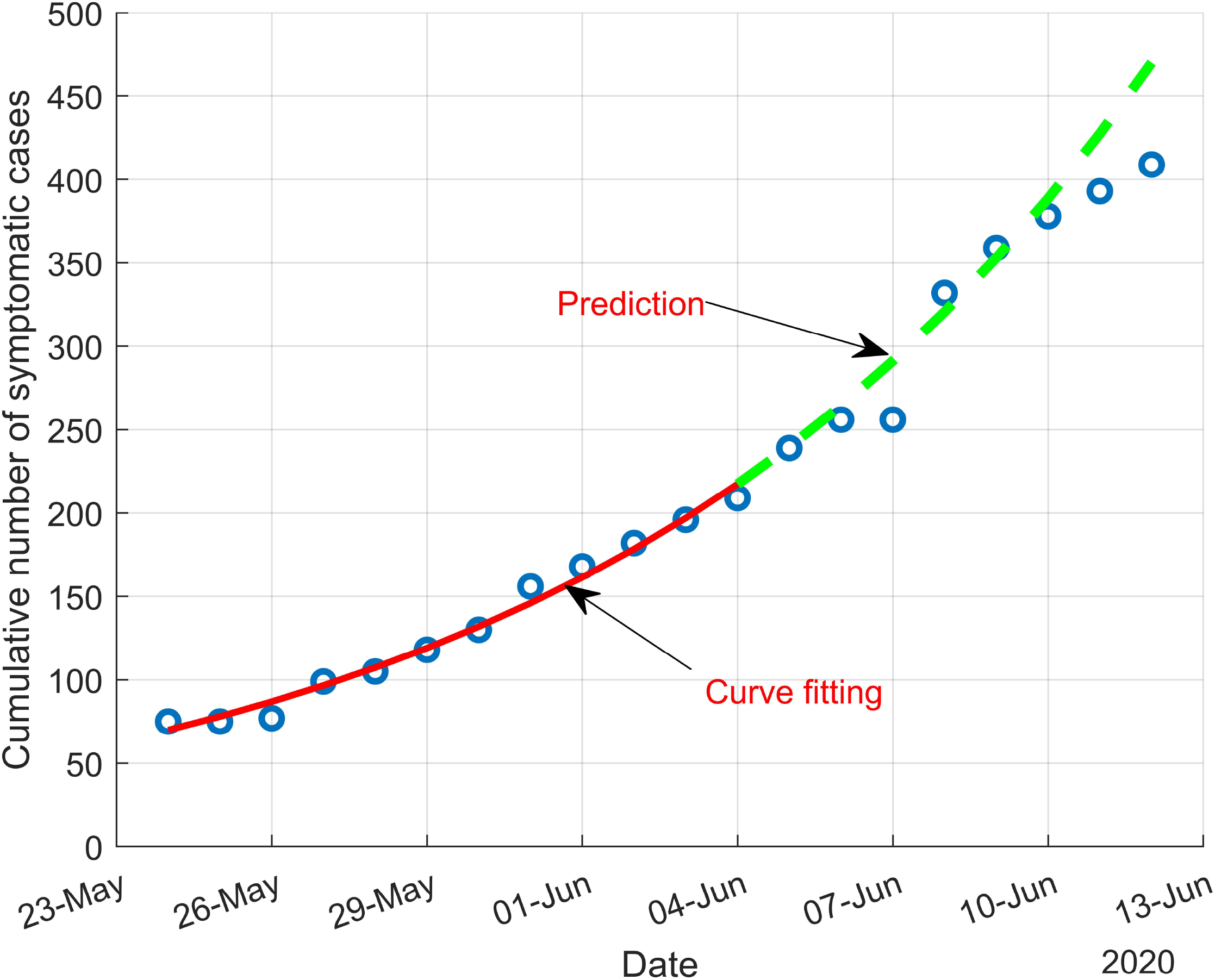

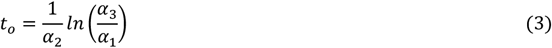

Applying equation (3) leads to *t*_*o*_ = −18.52 days which corresponds to May 5^th^.

The regression results were used to predict the progression of the epidemic in the future. Figure 3 shows our prediction of the outbreak compared to the actual data. Starting from June 4^th^, the prediction is up to 8 days in the future.

### The number of active cases

The distribution of the active cases can be estimated using equation (1c) in addition to nonlinear regression on the active cases data. First, we can write equation (1c) in the form

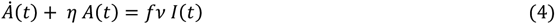

Substitute equations (2) and (1e) into equation (4) yields

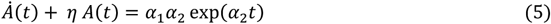

Equation(5) is a linear differential equation of the first order in the form 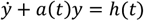 where *a*(*t*) *= η* and *h*(*t*) *= α*_1_*α*_2_ exp(*α*_2_*t*). This equation can easily be solved using an integration factor ^27^. The steps to solve this equation are outlined in Appendix C, and the final solution can be expressed as

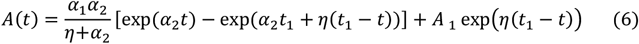

Where *t*_1_is the time immediately before the first infected individuals show symptoms and are hospitalized, and *A*_*1*_*is the* initial value of symptomatic cases. The solution to equation (6) needs an initial value, which can easily be expressed as *A*(*t*) *= 0 when t = t*_*1*_ which yields *A*_*1*_ = 0. Therefore, equation (6) can be rewritten as

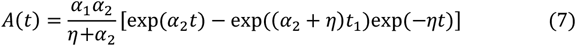

Equation (7) shows the distribution of the active cases, which can be rewritten as

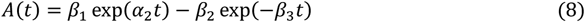

Where *α*_2_ is constant that was already evaluated from the cumulative number of cases as *α*_2_ = 0.0923 and β_*1*_, β_*2*_ and β_*3*_ are constants that can be expressed as

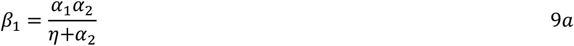

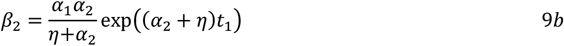

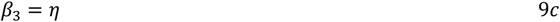

Equation (8) shows that the number of active cases have to follow two competing exponential distributions. The first exponential distribution (*β*_*1*_ exp(*α*_2_*t*)) is a measure of the number of newly added cases while the second distribution (−*β*_*2*_ exp(*−β*_*3*_*t*)) is a measure of how many active cases have been recovered by recovery or death.

The steps to evaluate *η* and *t*_1_, *β*_*1*_, *β*_*2*_ and *β*_3_ starts by fitting equation (7) to the known number of active cases from Table-A.2. The curve fit would yield *η* and *t*_1_, then using equations (9a -c) we could evaluate *β*_*1*_, *β*_*2*_ and *β*_*3*_.

Microsoft Excel solver ^28^ was used to fit equation (8) to the number of active cases from Table-A2. The solver used generalized reduced gradient method ^29^ to find optimum values for the parameters. The curve fit is shown in Figure 4, while the results for *η* and *t*_1_, were *η* = 0.02073, thus, the recovery period can be obtained as 1/*η* = 48 days, and *t*_1_ = −*3*.3 days which indicates that the first individuals started showing symptoms on May 21^st^ and from Figure 1 we could notice that around May 21^st^ is when the total number of symptomatic cases started to increase after it was relatively unchanged for almost a month.

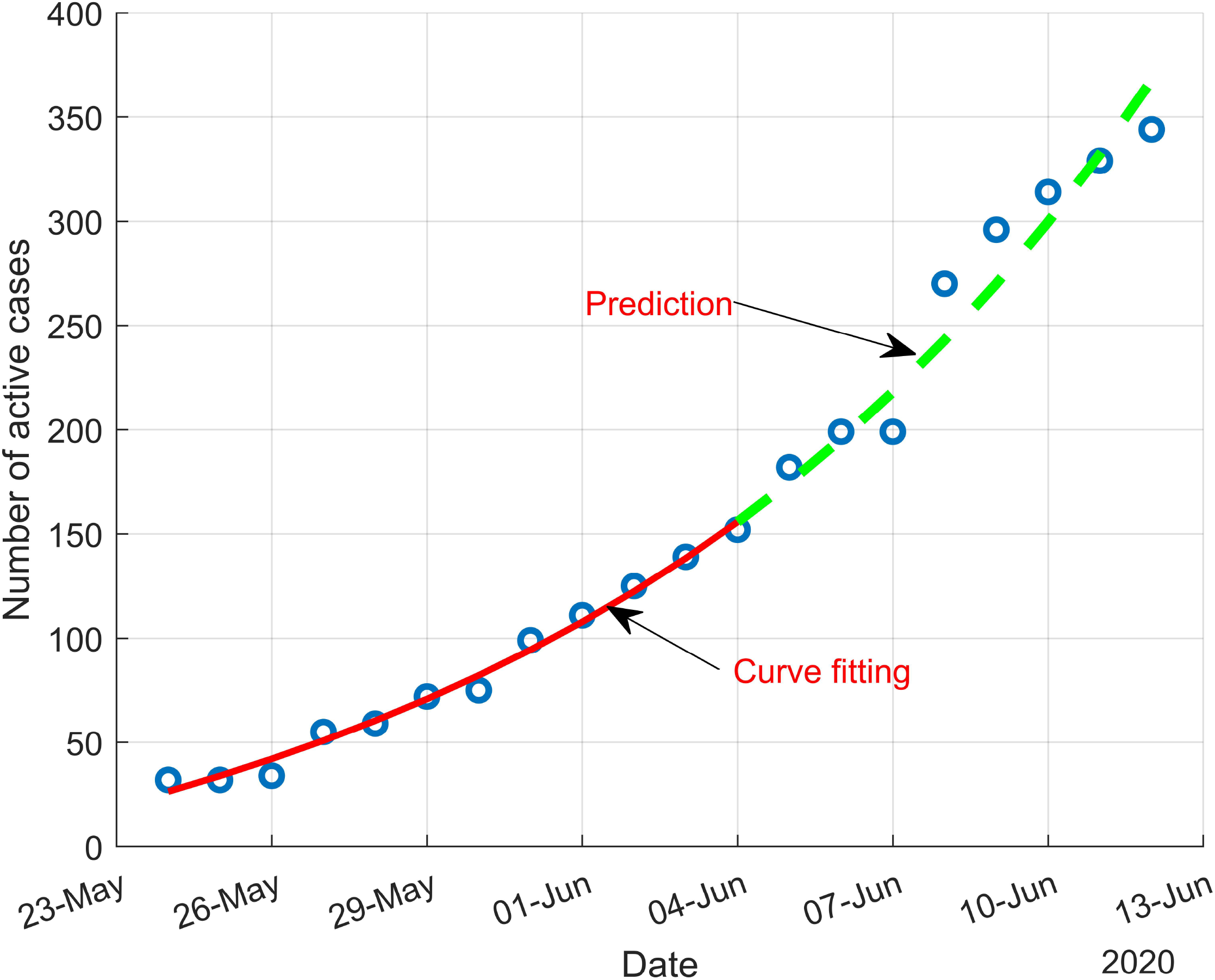

Equations (9a-c) are used to determine *β*_*1*_, *β*_2_ and *β*_*3*_ as 62.*3*39, 42.93 and 0.02073 respectively. Equation (8) with the known parameters *α*_*1*_, *β*_*1*_, *β*_*2*_ and β_*3*_ can be used to predict the number of active cases in the future. This number is really important to help health officials plan for the people who would need hospitalization or quarantine in the foreseeable future. Figure 4 shows the prediction vs. real data for 8 days in the future starting from June 4^th^. Our prediction agrees very well with the actual data.

The incubation period for the virus at the beginning of the pandemic can be defined as the period between when the epidemic started (*t*_*o*_) and when the first case showed symptoms of the disease (*t*_1_). Or

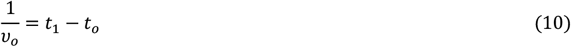

Applying equation (10) we get the initial incubation period as 1/*v_°_* = 15.2 days.

### Number of unreported cases

The number of unreported cases can be estimated from equation (1d). Equation (1d) can be rewritten in the form

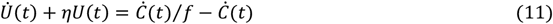

Substitute equation (2) and (1e) in equation (11) yields.

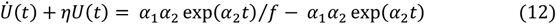

Equation(12) is a linear differential equation of the first order in the form 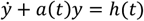 where a (*t*) *= η* and *h*(*t*) *= α*_1_*α*_2_ exp(*α*_2_*t*)/*f* − *α*_*1*_*α*_2_ exp(*α*_2_*t*). This equation can also be solved by using an integration factor ^27^ as shown in Appendix C. The final solution can be expressed as

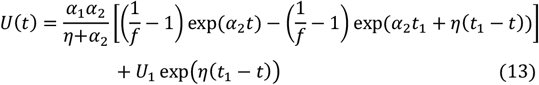

Equation (13) describes the change in the number of unreported cases with time. The only unknown in these equations are initial condition *U*_1_, and the percentage of the cases that become symptomatic *f*.

The initial value for the unreported cases can be safely assumed as *U*_1_ = *0*. Thus, equation (13) can be rewritten as

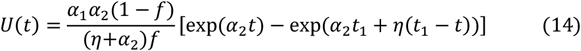

Again, we notice that there are two competing exponential terms in equation (14). The positive exponential term which is a measure of how many pre-symptomatic infected people become Asymptomatic and the negative exponential term which is a measure of how many Asymptomatic people recovered.

Until now, there’s no reliable study that gives information about the percentage of symptomatic cases from the total infected people, therefore, we could observe how the number of unreported cases change at different values of *f*, as shown in Figure 5

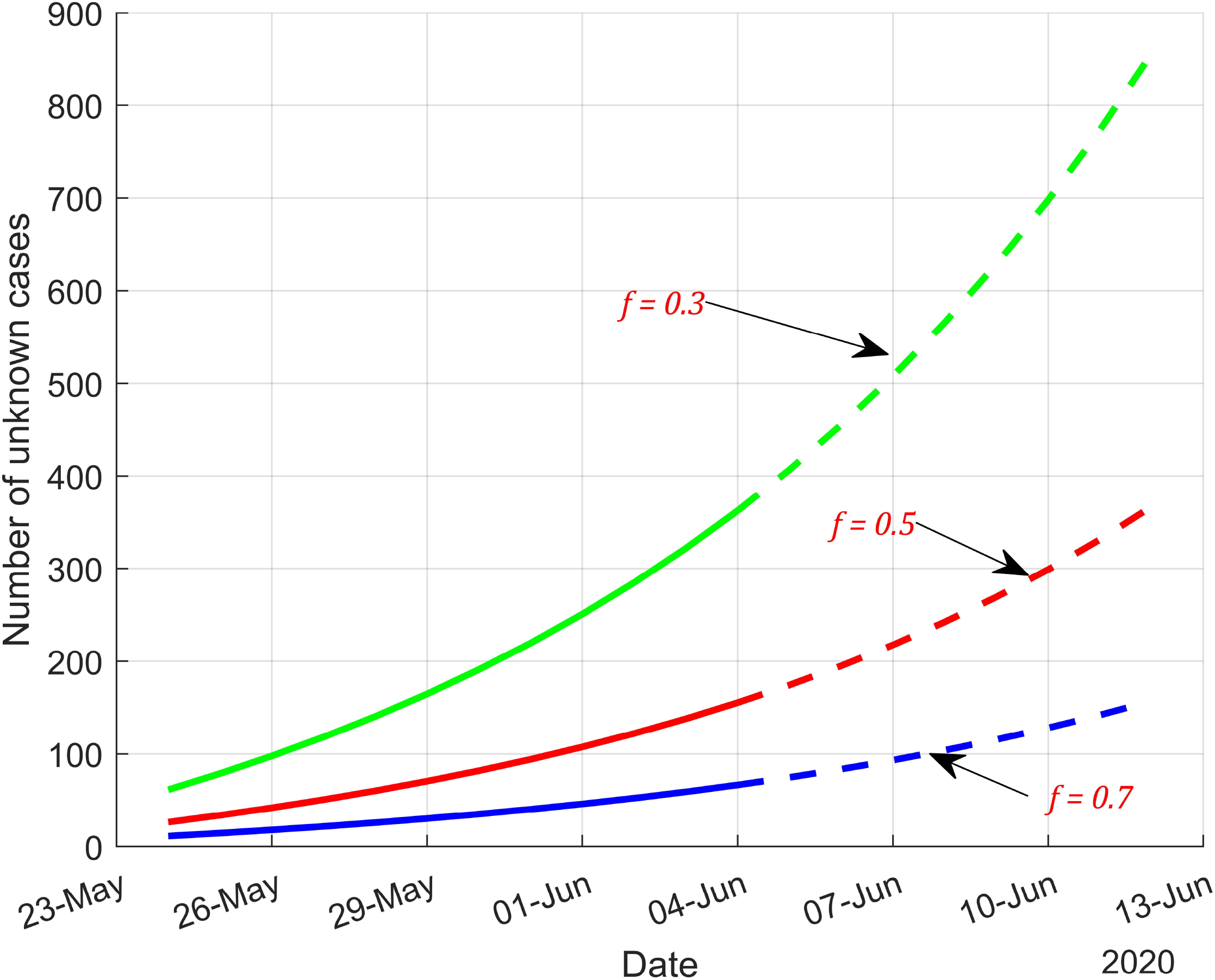

### Transmission rate and reproductive number

Now, let’s consider equation (1b). Equation (1b) is a nonlinear differential equation that describes the change in pre-symptomatic cases. Using, equation (1e) and (2) we can show that *İ = α*_2_*I*, then we substitute into equation (1b) and rearrange to get

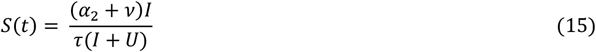

Now, we can apply the initial conditions, which are at *t*_*o*_ = *0, S = S*_*o*_, *I* = *I*_*o*_and *U* = *U*_*o*_ = 0, substitute for the initial conditions in equation (15) yields

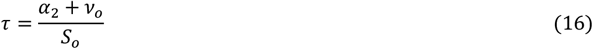

Equation (16) can be used to calculate the transmission rate, where *S*_*o*_is number of susceptible individuals at time *t*_*o*_. The COVID-19 virus affects everyone without discrimination, therefore in this case, the number of susceptible people is the entire population of Libya. The Libyan population is about 6.4 million, substituting for this number in equation (16) we get the transmission rate as *τ* = 2.47 × 10^−8^.

The reproductive number *R*_*o*_ is the number of the secondary cases generated by the primary cases at the start of the epidemic. It can be defined as the ratio between transmission rate per day times the number of susceptible individuals to the recovery rate ^3^. Or

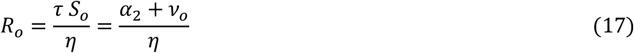

Using equation (17) we get the reproductive number as *R*_*o*_ *= 7* · 62.

## DISCUSSION

This research paper is the first that uses a new mathematical approach in order to calculate the parameters of the epidemiological model simply by finding a closed-form solution to the differential equations of the model without assumption of any parameter. However, for this approach to work, you need to have data that can easily be tracked which include the cumulative number of cases and the active number of cases.

The starting time of epidemic is calculated to be on May 5^th^, however, the first case was reported on March 24^th^. Indicating that the sudden increase in infectious cases was due to new source. This could be explained by the fact that the Libyan government allowed many citizens to return to the country starting from the beginning of May. The majority came from countries where the disease was highly prevalent like Turkey, Spain, Italy and Saudi Arabia; and some of them developed the disease shortly after arriving.

Our prediction of the cumulative number of symptomatic cases agrees very well with the actual data in the first 6 days but it over-predicted the number of cases for days 7 and 8 as shown in Figure 3. This could be attributed to the fact that the model parameters we found from the curve fitting are the average values for the period we covered (May 24^th^-June 4^th^), and these values usually change as people start taking more precautionary measures. Therefore, we recommend that the prediction process be performed for only one week and should be updated daily to account for any change in the model parameters due to the change in people’s social activities.

An exact solution to the number of active cases can be found mathematically as shown in equations (6-9). This solution was fitted to the data from Table-A.2, where we found that the period to recovery was 48 days. Also, the incubation period was found as 15.2 days, which is consistent with the available incubation period of the virus which is on average between 2-14 days and could reach up to 27 days ^30^. Please note, that 15.2 days are the incubation period at the start of the epidemic and not the average incubation period. Our prediction for the number of active cases is shown in Figure 4 agreeing very well with the actual data.

Similarly, an exact mathematical solution to the number of unknown cases can be obtained as shown in equations (13) and (14). However, there is no reliable way to know the percentage of infected people that become symptomatic (*f*). Therefore, we can’t estimate the number of unknown cases, but we could make an educated guess. Figure 5 shows the number of unknown cases at different values of *f*. Assuming that 70% of the infected individual become symptomatic cases (*f* = *0* · *7*) *th* en we expect the number of unreported cases to be around 157 individual by June/12 and this number increases to 366 individuals if *f* = *0* · 5 and to 855 cases if *f* = *0* · *3*.

The reproductive number was determined as 7.62 which is alarmingly high, and higher than that estimated in Wuhan, China before implementation of public health intervention measures^31^. The reproductive number depends on the social activities and the average number of people who became in contact with an infected person. Therefore, it is expected to vary from country to country. In general, Libyan people have large families and are socially active. The first cases were identified as people who returned home from abroad, and the customary social norm is that the friends and extended family visit a person who returns from travel. Therefore, we are expecting a large reproductive number at the early stages of the epidemic.

## CONCLUSION

With only the cumulative number of cases and the total number of active cases of COVID-19, assuming that the growth of cumulative number of cases follow an exponential growth, several important epidemic model parameters can be measured effectively. Even though this new mathematical approach is used to study the COVID-19 pandemic in Libya; it has the capability to be applied with any compartmental epidemic model of any infectious disease anywhere in the world.

## Data Availability

The data are available publicly

